# Predicting Vaping Cessation in Young Adults: A Machine Learning and Explainable Artificial Intelligence (XAI) Approach to Public Health Intervention

**DOI:** 10.1101/2025.09.15.25335817

**Authors:** Poolakkad S Satheeshkumar, Ian Lango, Swarnali Zafor, Mikaiel Ebanks, Rahul Kumar Das, Kit wai Cheung, Roberto Pili, Supriya D. Mahajan

## Abstract

The public health impact of vaping in the United States reflects a complex balance of potential benefits and emerging risks. While e-cigarettes can substantially reduce exposure to toxic combustion byproducts and may aid in smoking cessation for adult tobacco users, evidence links e-cigarette use to respiratory and cardiovascular injury, raising concerns about long-term health outcomes in vapers. Unfortunately, vaping has become deeply entrenched among youth. In 2024, 38.4% of adolescent e-cigarette users reported habitual vaping patterns, underscoring persistent nicotine dependence within this vulnerable population. This escalating youth uptake underscores the urgent need for comprehensive, evidence-based policies that simultaneously advance cessation support for adults and enable robust prevention strategies for adolescents. Thus, to inform the optimal use of predictive technologies in vaping cessation efforts, we conducted a social-media–based survey targeting young adult vapers. Our aims were to (1) characterize cessation-related behaviors and attitudes and (2) evaluate machine learning and XAI methods for predicting quit attempts and successes within this population In our study, we employed both forward selection and backward elimination techniques to identify key predictors of successful vaping cessation. The dataset was partitioned into training (70%) and testing (30%) subsets to facilitate model development and evaluation. We applied a range of machine learning algorithms to the training data and subsequently validated their performance on the test set. For linear modeling, we utilized least absolute shrinkage and selection operator (LASSO), ridge regression, and elastic net. In addition, we incorporated non-linear approaches including random forest (RF) and support vector machine (SVM) to capture more complex relationships within the data. We assessed the model performance through C-Statistics/ area under the curve (AUC). Further we validated the performance through Brier Scores. Among the models evaluated, linear approaches demonstrated superior overall performance, with non-linear models such as random forest (RF) and support vector machine (SVM) exhibiting strong predictive accuracy on the training data. LASSO regression yielded robust results, with area under the curve (AUC) values of 0.89 for the training set and 0.91 for the test set. Ridge regression followed closely, achieving AUCs of 0.88 and 0.93, respectively. Elastic net regression performed consistently across both datasets, with an AUC of 0.91 in training and testing.

Key predictors of successful vaping cessation included age, environmental triggers, vaping frequency, sex, and long-term behavioral outlook. Age emerged as a particularly influential factor, with individuals under 25 exhibiting increased vulnerability—likely due to neurodevelopmental sensitivity and elevated usage rates. Environmental cues, especially social exposure, were strongly associated with relapse risk, highlighting the importance of trigger management in cessation strategies. Interestingly, vaping frequency served as a counterintuitive indicator: erratic usage patterns correlated with lower cessation success, suggesting that consistent use may reflect a greater readiness for behavioral change. Sex-based differences were also notable, with males demonstrating more intense withdrawal symptoms and higher consumption levels, underscoring the need for gender-responsive interventions.

These findings underscore the utility of machine learning in uncovering nuanced behavioral and contextual determinants of addiction and cessation outcomes, offering valuable insights for the design of targeted public health interventions.

## INTRODUCTION

Over the past two decades, the use of electronic cigarettes (e-cigarettes) to inhale aerosolized nicotine and other substances, commonly referred to as vaping, has emerged as a pressing public health issue in the United States. Initially marketed as a harm-reduction strategy for adult smokers, e-cigarettes have rapidly gained traction among adolescents and young adults, driven by appealing flavors, aggressive promotional tactics, and perceptions of reduced health risk compared to combustible tobacco products [1].

Despite regulatory efforts, youth vaping remains a persistent concern. In 2024, approximately 1.63 million middle and high school students reported current e-cigarette use, representing a 5.9% prevalence rate. Although this marks a decline from the 2019 peak of 5 million users, the sustained levels underscore the challenge of curbing nicotine initiation among minors [2]. Among adults, the prevalence of vaping reached 6.9% in 2021, with the highest rates observed in individuals aged 18–24, reflecting a notable shift in nicotine consumption patterns [3, 4].

The effects of vaping in the United States are complex, to say the least. On one hand, public health viewpoints emphasize the potential of e-cigarettes to diminish exposure to harmful substances found in conventional tobacco, with research indicating that they may assist certain adult smokers in cessation efforts [5]. Nonetheless, apprehensions over long-term health repercussions—including pulmonary and cardiovascular hazards as evidenced in recent studies [6] and the elevated prevalence of habitual usage among adolescents (38.4% of 2024 users [2])— highlight considerable disadvantages.

The vaping industry is booming, as it is anticipated to generate a global market of $31.5 billion by 2029, all while consumers are incurring annual healthcare costs of approximately $2,024 per user. Socially, it has impacted tobacco consumption patterns, with adult cigarette smoking decreasing from 42% in the 1960s to 11.5% currently, which is partially attributable to the rise of vaping. However, this rapid increase in youth vaping has led to regulatory measures, including flavor bans and enforcement against illicit products, resulting in the seizure of 678,684 devices by October 2024. Measures including these highlight the conflict between harm reduction and steps already being taken for prevention. This evolving landscape has sparked ongoing debate regarding the dual role of e-cigarettes: as a potential aid in smoking cessation for established smokers and as a driver of nicotine dependence among non-smoking populations, particularly youth.

Vaping cessation has become a significant concern, with 63.9% of teenage vapers in 2020 indicating a desire to quit and 67.4% having attempted cessation in the previous year [10]. Text-based cessation programs have yielded promising results. Initiatives such as the Truth Initiative’s “This is Quitting” have proven quite effective, increasing cessation rates by 35-40% among young adults by providing automated text messages that give social support alongside giving cognitive and behavioral coping skills training [11]. Such interventions highlight the potential of scalable, technology-driven approaches to address vaping addiction. In this context, a critical question arises: Can predictive modeling serve as an optimal strategy for vaping cessation? Predictive algorithms, particularly those powered by machine learning (ML), offer the ability to identify individuals at higher risk of continued use or those predisposed to successful cessation.

Predictive algorithms can identify those predisposed to cessation or at risk of continuing usage in order to facilitate targeted interventions, which is particularly relevant given the 6.9% adult and 5.9% youth vaping prevalence rates in the United States [2, 3]. Machine learning and further Explainable Artificial Intelligence (XAI) improve predictions by offering explicit insights into the factors influencing cessation likelihood, hence enhancing trust and application in public health policies [12]. However, data constraints, implementation obstacles, and the demonstrated efficacy of prevention (e.g., the decline in youth vaping post-2019 [2]), direct support (e.g., cessation programs [11]), and extensive cessation research [13] indicate that prediction serves as a supportive tool rather than the most effective independent strategy [14].

As vaping evolves, comprehending its prevalence, effects, and cessation strategies, including the role of prediction, remains crucial for addressing its public health implications in the United States. To explore this further, data were collected via a social media-based survey to examine the characteristics associated with vaping cessation among young adults. The study employed ML and XAI techniques to identify predictive factors and interpret cessation outcomes, contributing to a nuanced understanding of how technology can support public health efforts in reducing e-cigarette use.

As vaping behaviors continue to evolve, it is imperative to maintain a multifaceted approach that integrates predictive analytics with proven cessation strategies. Doing so will enhance the effectiveness of public health interventions and ensure a more comprehensive response to the challenges posed by vaping among young adults. Understanding these dynamics is critical for informing public health policy, regulatory frameworks, and targeted intervention strategies.

## RESULTS

A total of 119 individuals participated in the study. The majority of respondents identified as Caucasian (58.5%), while the remaining 41.5% identified as African American, Hispanic, Asian, or other racial/ethnic backgrounds. In terms of age distribution, most participants were between 21 and 26 years old (74.6%), followed by 15–20-year-olds (16.1%). A smaller proportion (9.3%) fell into the combined age category of 27–32, 45–50, and 51–56 years. Female respondents comprised the majority of the sample (70.3%), with males accounting for 29.7%. No participants identified as another gender.

Regarding the age of vaping initiation, 11.0% of respondents reported starting at or before age 14, while 39.8% began between ages 15 and 18. Another 37.3% initiated vaping between ages 19 and 22, and 11.9% started at age 23 or older. Duration of vaping varied, with nearly half of respondents (49.6%) reporting use for more than four years. Additionally, 27.7% had vaped for three to four years, 15.1% for one to two years, and 7.6% for less than one year or did not respond.

Vaping frequency was notably high among participants. A majority (65.3%) reported vaping multiple times per day, exceeding five sessions daily. Others reported vaping once a week (21.2%), every other day (4.2%), or once daily (9.3%). Puff intensity also varied, with 55.1% of respondents taking more than 21 puffs per session. Smaller proportions reported taking fewer puffs: less than five (14.4%), six to ten (11.9%), eleven to fifteen (9.3%), and sixteen to twenty (9.3%).

Only 5.1% of respondents indicated that they vaped for weight loss purposes. When asked about desired effects, 38.1% reported vaping for the high or head rush, 29.7% for satisfaction, relaxation, or anxiety relief, and 18.6% for head effects alone. A further 13.6% cited a combination of head rush and emotional relief. Influences on vaping behavior were primarily personal (61.0%), followed by social (17.8%), other (11.9%), and habitual (9.3%).

Triggers for vaping included sensory cues, with 13.6% of respondents identifying smell as a primary trigger. Device preferences varied: 47.0% used rechargeable devices, while others used disposable devices with puff capacities of 1500 (14.5%), 2000 (8.5%), or 2500 (10.3%). Nearly one-fifth of participants (19.7%) did not respond to the device type question.

To evaluate predictors of vaping cessation, the following modeling approaches were employed - Lasso regression, Ridge regression, Random Forest, Elastic Net, and Support Vector Machine (SVM). Each model underwent hyperparameter tuning using cross-validation to identify the optimal regularization parameter (λ), followed by refinement of the decision rule.

### Statistical Modeling and Hyperparameter Optimization for Predicting Vaping Cessation

In the lasso, the lambda was optimized and used to create a model; after which the decision rule was optimized. The lambda values were selected from multiple values with the help of cross-validation. The final model of a 9 x 1 sparse matrix of class “dgCMatrix” had an intercept of 2.4640598 and coefficients of age (-0.8349198), sex (-1.0683805), vape age (-0.2031326), vape product (-0.3555436), type of interest in a vaping product (0.5456494), vape effects (0.9373031), vaping trigger (-0.7820531), and knowledge of vaping’s adverse effects (2.6913733). The lambda minimum of the final model was 0.00808717.

In the ridge, the lambda was optimized and used to create a model; after which the decision rule was optimized. The lambda values were selected from multiple values with the help of cross-validation. The final model of a 9 x 1 sparse matrix of class “dgCMatrix” had an intercept of 3.8464110 and coefficients of age (-0.8729834), sex (-0.6809932), vape age (-0.1592762), vape product (-0.2354564), type of interest in a vaping product (0.2640376), vape effects (0.4963433), vaping trigger (-0.6442088), and knowledge of vaping’s adverse effects (1.4071456). The lambda minimum of the final model was 0.03313344.

In the random forest, the lambda was optimized and used to create a model; after which the decision rule was optimized. The lambda values were selected from multiple values with the help of cross-validation. The final model had coefficients of age (3.923317), sex (1.844180), vape age (4.040729), vape product (5.738112), type of interest in a vaping product (2.624851), vape effects (2.667517), vaping trigger (3.572105), and knowledge of vaping’s adverse effects (2.243317).

Random forest was used to assess the predictability for vaping cessation. The mean decrease in Gini for age (3.923317), sex (1.844180), vape age (4.040729), vape product (5.738112), type of interest in a vaping product (2.624851), vape effects (2.667517), vaping trigger (3.572105), and knowledge of vaping’s adverse effects (2.243317) (Figure S3).

Cross-validation for lasso is provided in Supplementary Figure 1. In the plot, lambda demonstrates the tuning parameter: the 10-fold cross-validated binomial deviance as a function of (log) lambda (λ) for the lasso regularized model. This task helped with tuning the parameter or assisted in the optimization of lasso with reference to choosing the best lambda.

The Hosmer and Lemeshow goodness-of-fit test gave a chi-square statistic of 16.087 and a p-value of 0.04115 in the lasso training dataset and a chi-square statistic of 15.559 and a p-value of 0.04914 in the lasso test dataset. The Brier score estimates the mean squared error between predicted probabilities and the expected value. The calculated Brier score for the Lasso training model was 0.1134826. The calculated Brier score for the lasso test model was 0.1348965.

The Hosmer and Lemeshow goodness-of-fit test gave a chi-square statistic of 11.965 and a p-value of 0.1528 in the ridge training dataset and a chi-square statistic of 8.0108 and a p-value of 0.4324 in the ridge test dataset. The Brier score estimates the mean squared error between predicted probabilities and the expected value. The calculated brier score for the ridge training model was 0.1294758. The calculated brier score for the ridge test model was 0.08580645.

The Hosmer and Lemeshow goodness-of-fit test gave a chi-square statistic of 12.562 and a p-value of 0.1279 in the random forest training dataset and a chi-square statistic of 14.658 and a p-value of 0.06615 in the random forest test dataset. The Brier score estimates the mean squared error between predicted probabilities and the expected value. The calculated Brier score for the random forest training model was 0.04587181. The calculated Brier score for the random forest test model was 0.1723423. (See Table I)

**Table 1:** MODEL PERFORMANCE COMPARISON.

| Table 1: MODEL PERFORMANCE COMPARISON |  |  |  |  |
| --- | --- | --- | --- | --- |
| Model | Dataset | Hosmer & Lemeshow<br>$\chi^2$ squared | p-value | Brier Score |
| Lasso | Training | 16.087 | *0.04115 | 0.11348 |
|  | Test | 15.559 | *0.04914 | 0.13490 |
| Ridge | Training | 11.965 | 0.1528 | 0.12948 |
|  | Test | 8.0108 | 0.4324 | 0.08581 |
| Random Forest | Training | 12.562 | 0.1279 | 0.04587 |
|  | Test | 14.658 | 0.06615 | 0.17234 |

To interpret the influence of individual predictors on the model’s output while accounting for feature interactions and correlations, Accumulated Local Effects (ALE) plots were employed. ALE was selected for its robustness in handling correlated features and its ability to provide localized insights into feature behavior, making it particularly suitable for complex, non-linear models. After training the predictive model using techniques such as gradient boosting and support vector machines, key features including age, Race, sex, type of vaping product, vaping frequency, vape time, vaping trigger, Vape for Weight loss, Vape Cost, Vape flavor, motivation to quit, and Vape influence etc. were selected for interpretability analysis (Figure 3). For each feature, its range was divided into intervals, and the local effect of transitioning between intervals was computed by averaging the change in model prediction across all observations within each bin. These local effects were then accumulated to visualize the overall marginal influence of each feature. ALE plots, with visualizations displaying feature values on the x-axis and the accumulated effect on the y-axis (Figure 3). This approach allowed for a nuanced understanding of how each predictor contributed to the likelihood of successful vaping cessation, while mitigating the distortions often introduced by feature correlation in other interpretability methods such as partial dependence plots.

**Figure 1:**
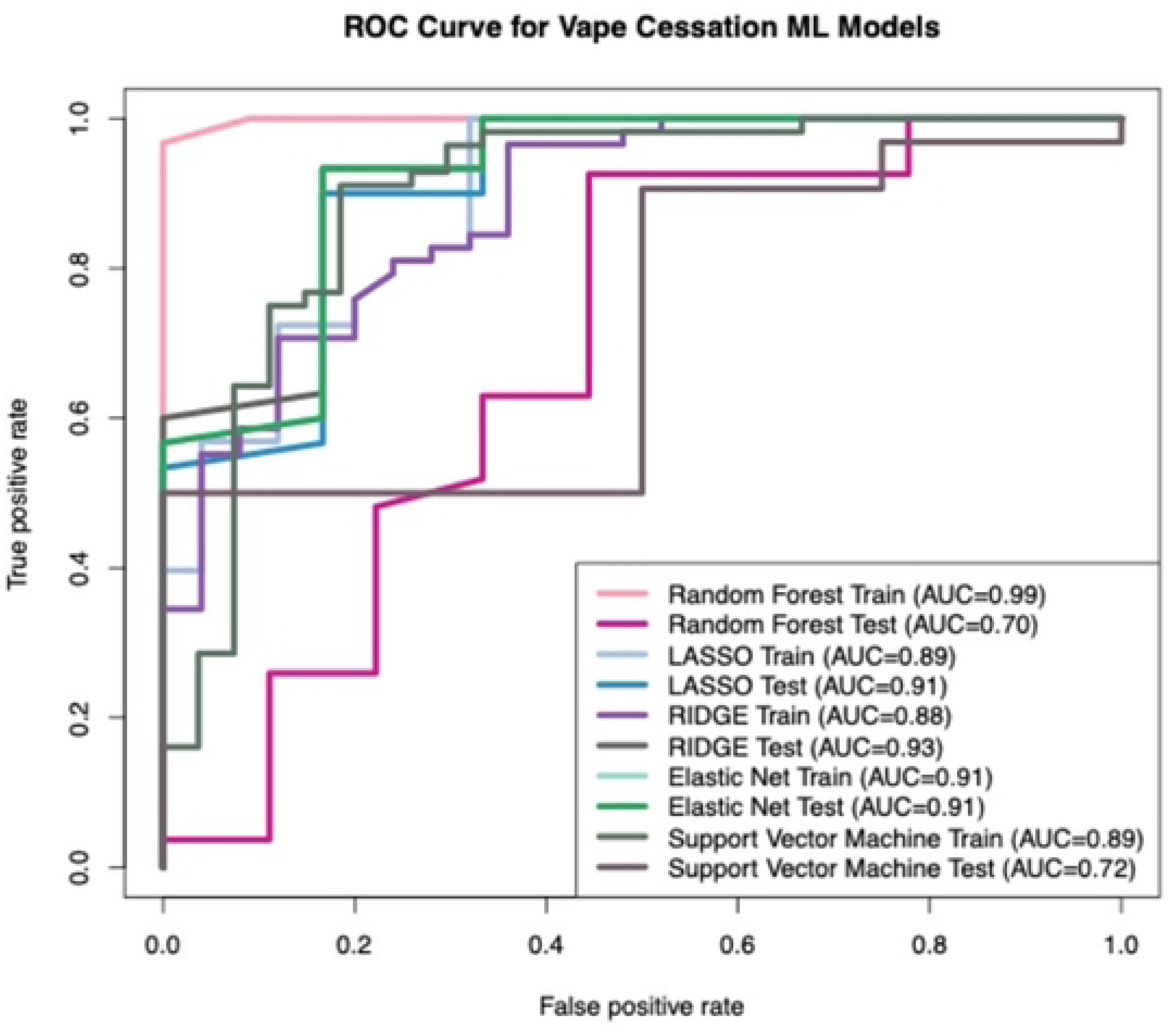
Receiver Operating Characteristic (ROC) Curves for VapeQuit Machine Learning Models: This figure displays the performance of various machine learning models (Random Forest, LASSO, RIDGE Elastic Net and Support Vector Machine) for this study, illustrating their ability to distinguish between positive and negative cases. Each curve represents the trade-off between the True Positive Rate (Sensitivity) on the y-axis and the False Positive Rate (1 - Specificity) on the x-axis across different classification thresholds. The Area Under the Curve (AUC) for each model’s training and testing phases is provided below:

- Random Forest Train (AUC=0.99): Represents the performance of the Random Forest model on the training dataset.
- Random Forest Test (AUC=0.70): Represents the performance of the Random Forest model on the independent testing dataset.
- LASSO Train (AUC=0.89): Represents the performance of the LASSO model on the training dataset.
- LASSO Test (AUC=0.91): Represents the performance of the LASSO model on the independent testing dataset.
- RIDGE Train (AUC=0.88): Represents the performance of the RIDGE model on the training dataset.
- RIDGE Test (AUC=0.93): Represents the performance of the RIDGE model on the independent testing dataset.
- Elastic Net Train (AUC=0.91): Represents the performance of the Elastic Net model on the training dataset.
- Elastic Net Test (AUC=0.91): Represents the performance of the Elastic Net model on the independent testing dataset.
- Support Vector Machine Train (AUC=0.89): Represents the performance of the Support Vector Machine model on the training dataset.
- Support Vector Machine Test (AUC=0.72): Represents the performance of the Support Vector Machine model on the independent testing dataset.

**Figure 2:**
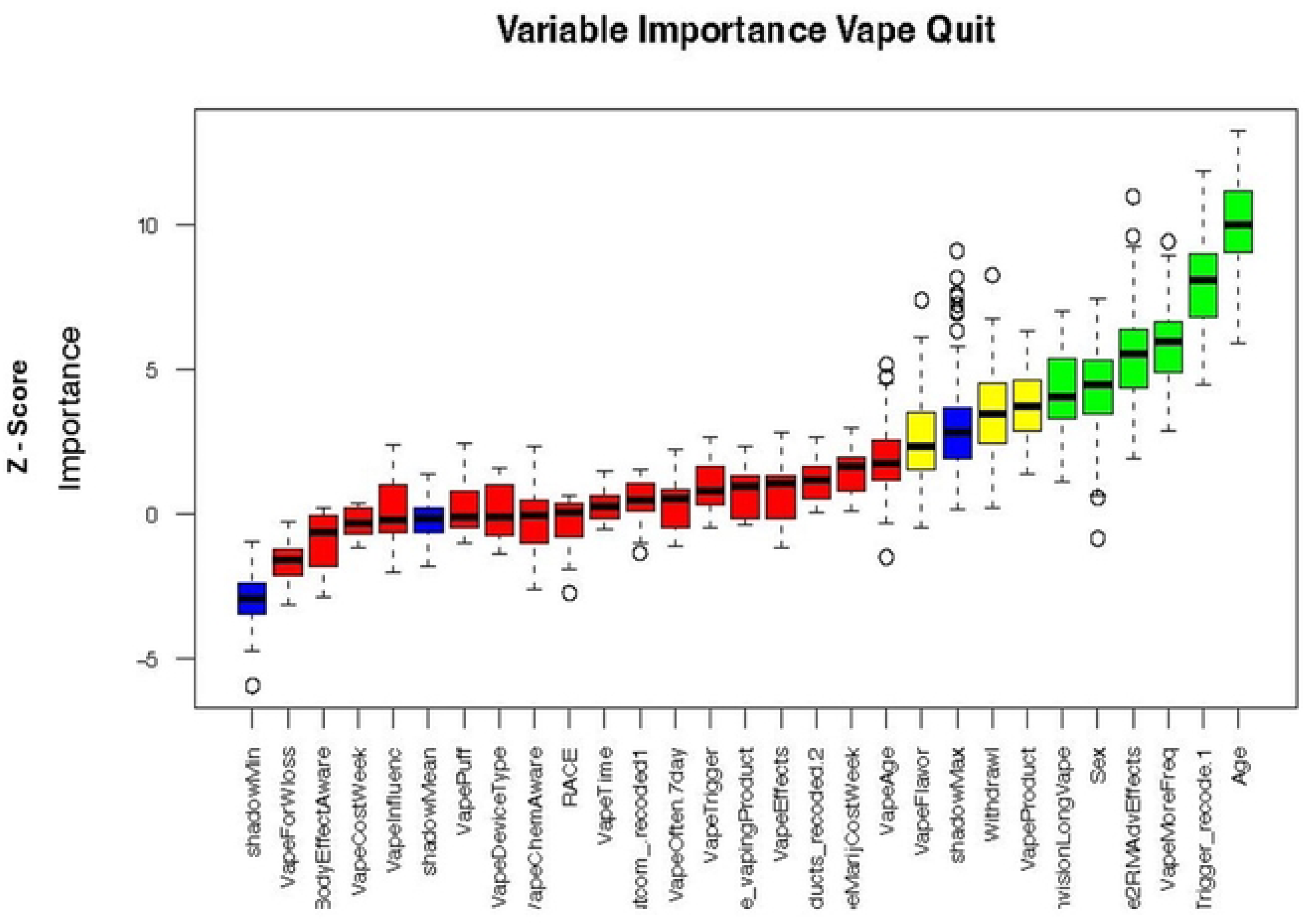
Variable Importance Plot for Predicting Vape Quit Behavior using the Boruta Algorithm. This box plot displays the importance of various variables in predicting vape quit behavior, as determined by the Boruta feature selection algorithm. The y-axis represents the Z-score, indicating the importance of each variable relative to shadow features (randomly permuted copies of original features). Interpretation of Boxplots:

- Blue boxplots: Represent the minimum, average, and maximum Z-scores of the shadow features, serving as a baseline for importance.
- Red boxplots: Indicate rejected variables, meaning their importance (Z-score) is consistently lower than the maximum Z-score of the shadow features, suggesting they are unimportant for predicting vape quit.
- Yellow boxplots: Represent tentative variables, whose importance is inconclusive and requires further evaluation (not clearly rejected or confirmed).
- Green boxplots: Show confirmed variables, indicating their importance is statistically significantly higher than the shadow features, thus confirmed as important predictors of vape quit. The box plot visually distinguishes between confirmed important variables (e.g., Age, VapeMoreFreq, Sex) and rejected variables (e.g., VapeForWloss, BodyEffectAware), providing insights into factors influencing vape cessation.

**Figure 3:**
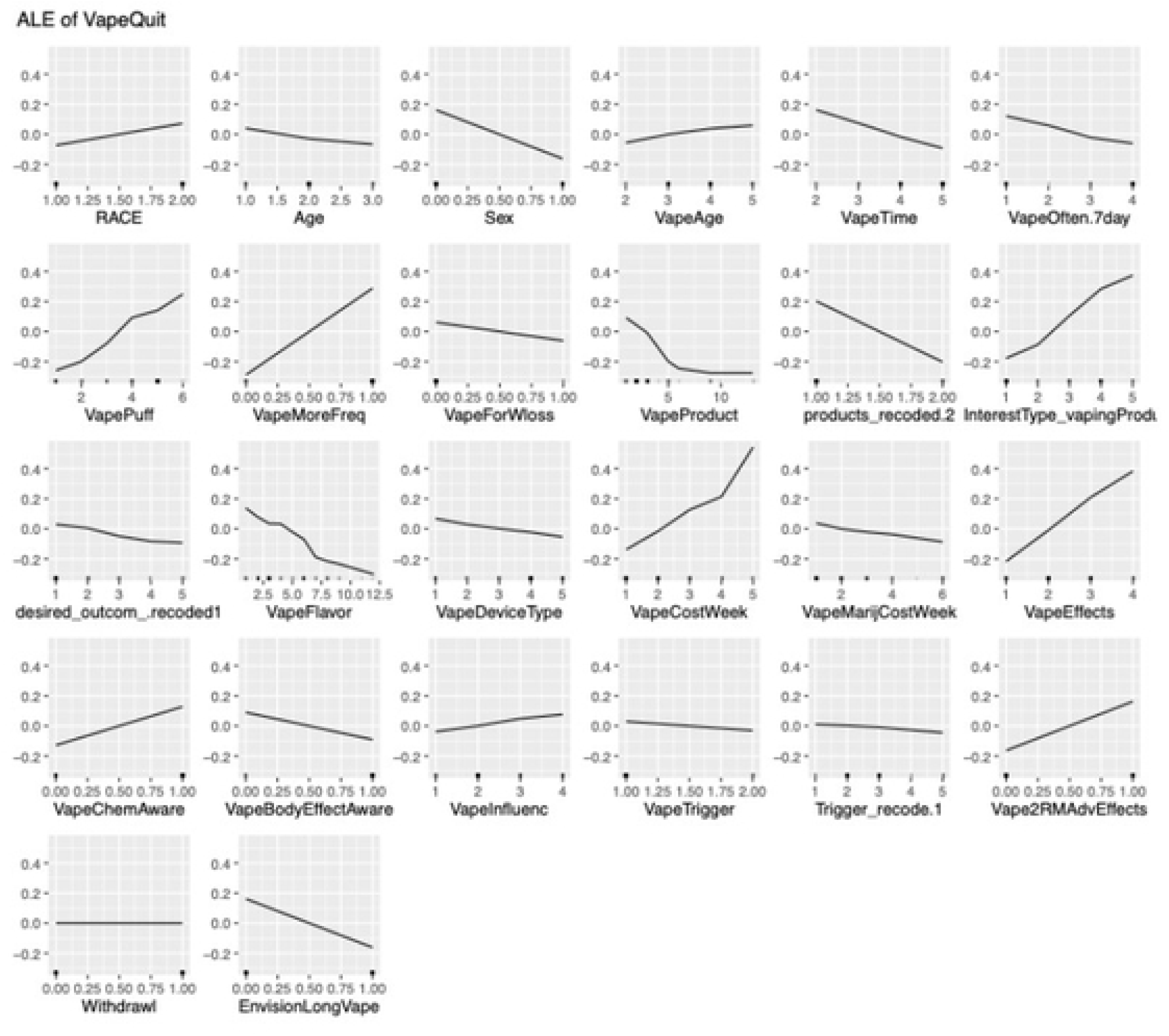
Accumulated Local Effects (ALE) plots. ALE plots enable interpretation of the influence of individual predictors on the model’s output while accounting for feature interactions and correlations. Figure X. Accumulated Local Effects (ALE) plots illustrating the marginal influence of key predictors on the likelihood of successful vaping cessation. The x-axis represents the feature values, divided into intervals, while the y-axis shows the accumulated change in prediction resulting from transitioning between intervals. ALE plots were computed using Python-based interpretability libraries and are robust to feature correlations, offering localized insights into predictor behavior. Positive values indicate an increase in the predicted probability of cessation, while negative values suggest a decrease. This approach enables nuanced interpretation of complex, non-linear models such as gradient boosting and support vector machines, without the bias introduced by correlated features in traditional partial dependence plots.

Random Forest XAI-based explainability was constructed using 83 samples (unweighted), 26 predicators, and two classes of “No” and “Yes.” (Figure 4). Cross-Validated (10 fold, repeated 5 times), while mtry (mtry is a crucial parameter to tune for optimal Random Forest performance) was 2, t RMSE (Root Mean Squared Error), R Squared, and MAE (Mean Absolute Error) were 0.401281, 0.2944698, and 0.3514571; while the mtry was 14, the RMSE, R Squared and MAE were 0.399175, 0.2686559, and 0.3303714; while mtry was 26, the RMSE, R Squared, and MAE were 0.397605, 0.2805989, and 0.3198172.

**Figure 4:**
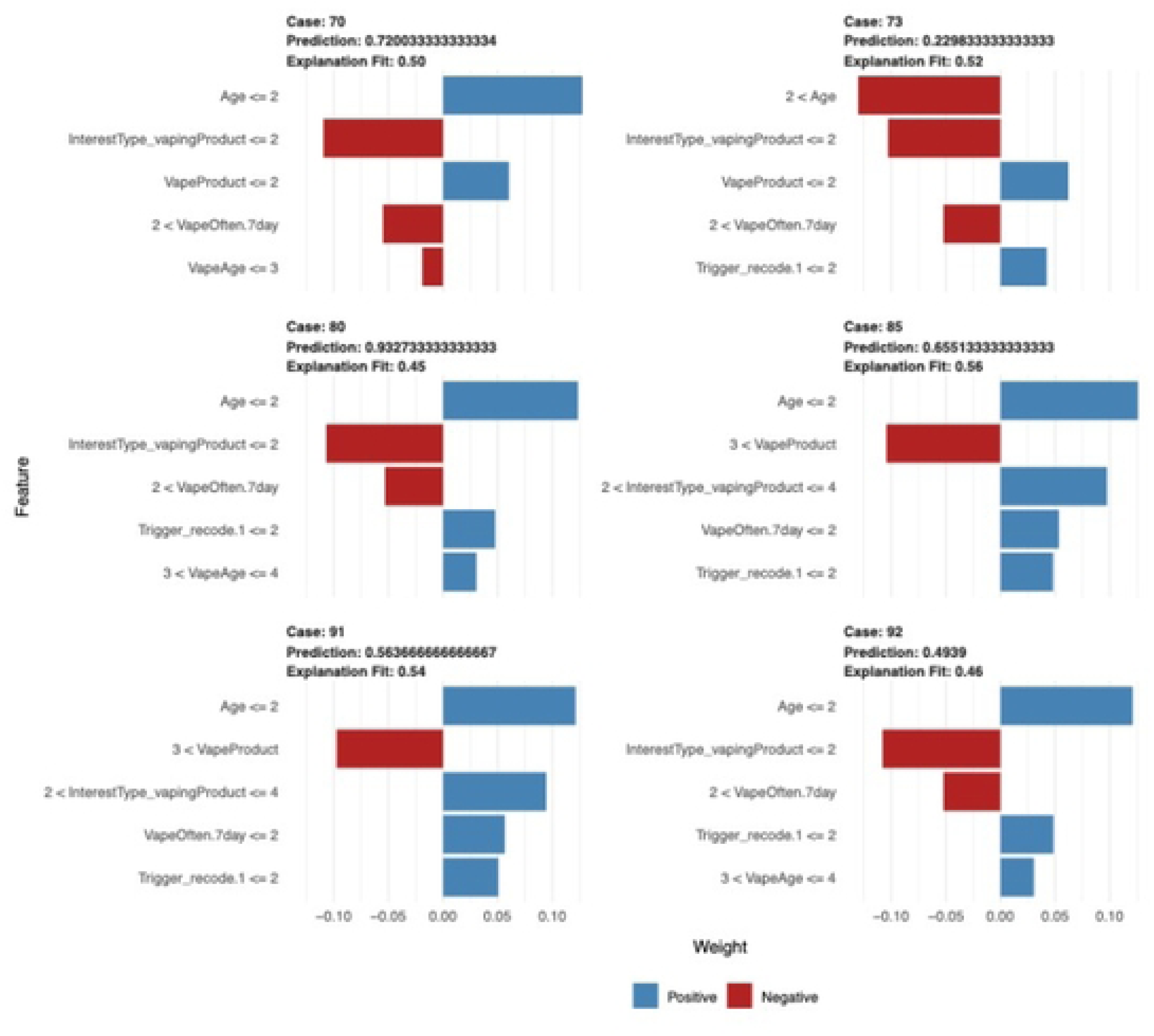
XAI (Explainable Artificial Intelligence) Based plot. Random Forest XAI-based explainability was constructed using 83 samples (unweighted), 6 predicators, and two classes of “Positive” and “Negative” influences.

The explanation plot shows the cases from 70 to 92, with predicted root variables transferred in and elective admission along with age, type of vaping product, vaping frequency and vaping trigger variables. The predicted probability ranged from 0.72 (case 70) to 0.49 (Case 92) with an explanation fit of 0.50 (case 70) to 0.46 (case 92) (Figure 4).

The Random Forest model demonstrated strong predictive capability and highlighted the relative importance of behavioral and knowledge-based factors in vaping cessation. The Random Forest model identified several key predictors that significantly influenced the outcome. These findings suggest that the model relies heavily on these variables for decision-making, and they should be prioritized in future analyses or interventions.

The Elastic Net model was optimized through cross-validation to balance variable selection and shrinkage. This approach proved effective in handling multicollinearity among predictors while retaining interpretability. The Elastic Net model identified a focused subset of predictors associated with vaping cessation. Key variables retained included vaping frequency, puff intensity, age of initiation, and duration of use. Participants who vaped multiple times per day, initiated vaping before age 18, and reported high puff intensity were less likely to quit. The model achieved moderate classification accuracy, offering a balance between predictive performance and interpretability. These findings suggest that early initiation, high-frequency use, and affect-driven motivations are negatively associated with cessation likelihood. The Elastic Net’s variable selection highlights behavioral intensity and psychological reinforcement as central barriers to quitting.

The Support Vector Machine (SVM) model, trained with a radial basis function kernel and tuned for optimal hyperparameters, demonstrated superior classification accuracy compared to linear models. SVM achieved high classification accuracy and strong discriminative power, outperforming linear models in terms of sensitivity and AUC. It effectively captured non-linear relationships among predictors. Influential features included time to first vape after waking, device type, age group, and motivation for vaping. Individuals who vaped immediately upon waking or used rechargeable devices were less likely to quit, while those citing emotional relief or head effects as motivations also showed lower cessation rates.

## DISCUSSION

### Effectiveness of Predictive Methods and Key Variables in Vaping Cessation

This study explored behavioral, demographic, and psychosocial predictors of vaping cessation among a diverse sample of 119 individuals, using five distinct modeling approaches: Lasso regression, Ridge regression, Random Forest, Elastic Net, and Support Vector Machine (SVM). Each model offered unique insight into the factors influencing cessation, shaped by its underlying assumptions and strengths. This study employed both forward selection and backward elimination techniques to identify key predictors associated with successful vaping cessation. These statistical approaches enabled the isolation of variables that significantly increased the likelihood of quitting vaping. Among the predictive models evaluated, nonlinear algorithms, particularly random forest, demonstrated superior performance, while linear models such as Lasso regression also yielded strong predictive capabilities. Lasso regression identified a sparse set of predictors most strongly associated with cessation. The model emphasized behavioral intensity, such as high puff frequency and early initiation, as key barriers to quitting. Its simplicity and interpretability make Lasso particularly useful for identifying actionable targets in intervention design. However, its tendency to exclude correlated variables may have limited its ability to capture the full complexity of vaping behavior. Ridge regression retained all predictors, shrinking coefficients to mitigate overfitting. While the Ridge model offered stable estimates, its inability to perform variable selection made interpretation more challenging. Ridge was less effective in isolating dominant predictors, especially in the presence of multicollinearity among behavioral variables such as frequency, puff intensity, and duration. Nonetheless, it provided a useful baseline for comparison and highlighted the cumulative influence of multiple low-impact features. Random Forest excelled in capturing non-linear relationships and interactions among predictors, and it identified vaping frequency, device type, and emotional motivations as top contributors to cessation outcomes. The model’s robustness and high predictive accuracy underscore the importance of complex behavioral patterns and contextual influences. Elastic Net model’s strength lies in its ability to handle multicollinearity and perform variable selection simultaneously. In this study, it highlighted behavioral intensity and early initiation as strong barriers to cessation. These findings align with existing literature suggesting that entrenched habits and early exposure to vaping are difficult to reverse. The model’s interpretability makes it valuable for informing targeted interventions, such as early prevention programs and behavioral counseling for high-frequency users. SVM’s ability to model complex, non-linear interactions revealed nuanced behavioral patterns not captured by Elastic Net. The model underscored the role of dependence indicators in predicting cessation outcomes. These insights suggest that cessation strategies should address both psychological triggers and product design. While SVM lacks the transparency of regression-based models, its predictive strength makes it a powerful tool for identifying high-risk individuals and tailoring interventions accordingly.

These findings underscore the utility of machine learning in public health research, particularly in identifying individual and contextual factors that influence addiction trajectories and cessation outcomes. The most salient predictors of vaping cessation included age, environmental triggers, vaping frequency, sex, and long-term envisionment of vaping behavior. These variables not only enhanced model accuracy but also offer actionable insights for tailoring cessation interventions and are discussed below.

### Age as a Determinant of Vaping Cessation

Age emerged as a critical variable, consistent with existing literature indicating that earlier initiation of nicotine use correlates with greater difficulty in cessation later in life. Individuals under 25 are particularly vulnerable due to ongoing neurodevelopment, which heightens susceptibility to nicotine addiction [18]. National survey data further support this, with approximately 20% of individuals aged 18–24 reporting e-cigarette use, and significant prevalence observed among middle and high school students (16.5% of 8th graders and 35.5% of 12th graders in 2020) [23,25]. Age also influences nicotine metabolism and vaping frequency, with emerging adults tending to vape more frequently than adolescents [16,18]. These age-related differences reinforce the importance of age-specific cessation strategies.

### Environmental Triggers and Relapse Risk

Environmental cues, such as vaping in social settings or exposure to others who vape, were strongly associated with continued use and relapse. These triggers can elicit cravings even among individuals actively attempting to quit. Evidence suggests that individuals who achieve long-term abstinence report fewer relapse triggers, highlighting the importance of trigger identification and management in cessation programs [15, 21]. Public health interventions should consider modifying environments and reducing exposure to common triggers to support sustained cessation.

### Vaping Frequency and Behavioral Stability

Vaping frequency was another robust predictor of cessation outcomes. Individuals with unstable vaping patterns were found to be 47% less likely to quit compared to daily users, suggesting that consistent use may paradoxically reflect greater behavioral awareness or readiness for change [17, 21]. Intervention efficacy has also been shown to vary by frequency, indicating that cessation programs should be calibrated to account for usage patterns and behavioral stability.

### Sex-Based Differences in Cessation Dynamics

Sex differences in nicotine reinforcement and withdrawal responses were evident, with male subjects exhibiting greater anxiety-like behaviors during withdrawal in preclinical studies. Epidemiological data further reveal that males tend to vape more frequently and engage in more episodes per day than females [24,25]. These physiological and behavioral differences necessitate sex-specific approaches to cessation, including tailored messaging and support mechanisms.

### Implications for Public Health Practice

The identification of these key variables through predictive modeling offers valuable direction for enhancing vaping cessation interventions. Machine learning tools can effectively stratify individuals based on risk and responsiveness, enabling more personalized and effective public health strategies. By integrating these predictors into clinical and community-based programs, practitioners can improve cessation outcomes and reduce the burden of nicotine addiction across diverse populations. Our findings will guide the integration of predictive analytics into comprehensive vaping cessation frameworks.

The use of multiple modeling approaches enriched the analysis, revealing both linear and non-linear relationships. While simpler models like Lasso and Elastic Net offer clarity and interpretability, machine learning methods such as Random Forest and SVM provide deeper insights into complex behavioral interactions. Together, these models offer a comprehensive framework for understanding and addressing vaping cessation.

The integration of predictive analytics and machine learning algorithms into public health strategies presents a promising frontier in addressing vaping-related nicotine addiction. Our study reinforces the utility of models such as LASSO and random forest in accurately identifying individuals at heightened risk for addiction, cessation failure, and relapse. Through our analysis, we were able to isolate critical behavioral and contextual factors, such as frequency of vaping and exposure to specific triggers, that contribute to these outcomes. These insights pave the way for more precise, data-informed intervention strategies tailored to individual risk profiles. Importantly, predictive methodologies not only enhance early identification and prevention efforts but also offer a framework for personalized cessation support and post-cessation relapse mitigation [18]. Their demonstrated success in tobacco cessation contexts underscores their potential for broader application in vaping-related care. However, despite their proven efficacy, these tools remain underutilized at both individual and population levels.

To maximize their impact, public health systems must prioritize the integration of predictive analytics into clinical workflows, community outreach, and policy development. Doing so will allow for more equitable, proactive, and effective responses to the evolving challenges of nicotine addiction. As vaping continues to rise among vulnerable populations, leveraging machine learning tools with help inform care standards.

By identifying key factors that predict vaping cessation, predictive analysis offers a powerful means to uncover and address underlying disparities that contribute to nicotine addiction. Previous research has demonstrated its effectiveness in improving patient outcomes in tobacco cessation by isolating actionable variables and informing targeted interventions [26]. Applying these same analytical techniques to vaping can similarly reduce negative health outcomes and enhance cessation success.

Beyond initial risk assessment and intervention planning, predictive models have also proven valuable in identifying individuals at heightened risk of relapse following smoking cessation [20]. By recognizing relapse-prone patterns, these tools enable the development of sustained support strategies tailored to vulnerable populations, ensuring long-term success beyond the point of initial cessation. The demonstrated utility of predictive analytics in tobacco-related contexts underscores the importance of extending these methods to vaping. Doing so can refine intervention efforts, personalize care, and ultimately improve outcomes for individuals struggling with nicotine dependence.

The demonstrated success of predictive methods in addressing smoking and tobacco addiction underscores their potential for broader application in the context of vaping. These analytical tools have proven effective in forecasting outcomes and identifying high-risk variables, making them invaluable for guiding intervention strategies. Despite their promise, predictive analysis remains underutilized at both individual and population levels, limiting its impact on care delivery and public health outcomes [19].

To bridge this gap, efforts must focus on integrating predictive models into clinical and community-based frameworks. By identifying key variables, and combinations of variables, that elevate an individual’s risk for vaping addiction or hinder cessation, these tools can inform more personalized and proactive approaches to care. The scientific consensus affirms the value of predictive analytics, and their strategic implementation can significantly enhance our understanding of vaping behaviors, improve treatment efficacy, and elevate care standards for affected populations.

Machine learning and predictive analysis are not only innovative but are also essential instruments for advancing public health responses to the growing challenge of nicotine dependence through vaping. Overall, predictive modeling offers actionable insights for public health practice. By integrating these variables into clinical and community strategies, machine learning can enhance intervention precision, improve cessation outcomes, and reduce nicotine dependence across diverse populations.

## MATERIALS & METHODS

### ETHICS STATEMENT

Our study was reviewed by University at Buffalo Institutional Review Board (UBIRB); Office of Research Compliance, Clinical and Translational Research Center Room 5018; 875 Ellicott St., Approval number: UBIRB IRB ID#: STUDY00005954; IRB Approval Statement - “The study materials for the project referenced above were reviewed and approved by the SUNY University at Buffalo IRB (UBIRB) by Non-Committee Review. The UBIRB has determined on 1/27/2022 that the research is Exempt according to 45 CFR Part 46.104. There is no expiration date”.

Survey was anonymous and voluntary. No personal information was obtained from the subjects who consented to participate. No children participated in the survey. Only adults >18 years of age participated in the survey.

### Data collection

An online anonymous survey was formulated to collect data on demographic information (age, gender, race/ethnicity), the individual’s vaping status, vaping frequency per day/week, the age when individuals started vaping, vaping duration from the time the individual started ever, which vaping products that the individuals use, questions pertaining to factors that contribute to individuals vaping, if individuals have ever had any adverse experiences while vaping, and if individuals ever tried to quit vaping, etc. Individuals were recruited across various social media platforms such as Snapchat, Instagram, Facebook, and Reddit Forum via direct messaging to potential participants and the link to the vaping survey questionnaire that was developed was distributed to participants online. This data was collected over a span of a six-week time period. There were 121 participants in the survey however, 10 participants were excluded due to those individuals not completing the survey in its entirety.

### Eligibility criteria

Individuals were eligible to participate in the study if they had a history of vaping in the past or if they currently used any vaping devices (nicotine vapes, tetrahydrocannabinol (THC) vapes, and cannabidiol (CBD) vapes).

### Statistical analysis, selection of variables, preprocessing, and ML methods

A chi-square test was used for the categorical variables and a t-test for continuous variables to demonstrate the baseline characteristics to stratify the groups between those with vape cessation vs those without vape cessation. The first objective was to fit the best subsets-feature selection in order to build an ML model. Forward selection and backward elimination regression methods were used for subset selection, with the best subsets being determined by minimum Akaike information criteria (AIC) from the forward selection and backward elimination. ML models were built using variables from forward selection and backward elimination. Linear and non-linear ML methods were used to build models. Linear models used least absolute shrinkage and selection operator (lasso). The lasso regression model was cross-validated, and performance was assessed in training and test sets. The model was trained on 70% training data and 30% test data. The regularization parameter for lasso was selected by tailoring for a considerable range of value of lambda and measured for the best value of lambda. Model performance by discrimination (C-statistics) and calibration was determined by using receiver operator curves (ROC) and Hosmer-Lemeshow test on training and test data. ML models were further assessed via stratification based on gender and then evaluated based on whether the developed methods performed better on males than females or vice-versa. All statistical analyses were performed with R 3.5.1 (R Foundation for Statistical Computing, Vienna, Austria).

### Prediction models and risk factor analysis

For subset selection in the construction of the ML model, forward selection and backward elimination regression techniques were used, with the minimal Akaike information criterion (AIC) used to determine the best subsets. Important variables were identified through forward selection and backward removal, with the results being used to construct ML models. The Boruta package was used to obtain and compare the feature selection models. Features that optimize model performance were used for building the ML model.

Both linear and non-linear ML techniques were used to construct the models. Least absolute shrinkage and selection operator (Lasso) ridge, and elastic net regression were used as the linear model, and RF was used as the non-linear model. Both the training set and test set were used to evaluate lasso regression model efficacy via cross-validation. The model was trained with 70% of the data and validated with the remaining 30%. Receiver operator curves (ROCs) and the Hosmer-Lemeshow test were used to compare the model’s performance on both the training and test sets. The same processes were repeated for the ridge and. Elastic Net (EN) regression. Elastic Net is a linear regression model that combines L1 (Lasso) and L2 (Ridge) regularization. Random forest (RF) was used to estimate ML model prediction and explainability (Supplementary Materials). Classification accuracy was measured using ROCs, Hosmer-Lemeshow test, and the confusion matrix.

RF was used for employing the explainable artificial intelligence (XAI); the process parameter tuning was included with elaborate computation through a parallel core through the use of “detectCores,” which were utilized to automatically determine the number of cores for “makePSOCKcluster” functions. Local Interpretable Model-Agnostic Explanations (LIME) techniques were used to achieve a means of local interpretation. R 4.6.2 was used for all statistical testing (R Foundation for Statistical Computing, Vienna, Austria).

## Data Availability

Data will be available on request

## ACKNOWLEGMENTS

The authors wish to express their sincere appreciation to all individuals who participated in this survey. Your time, insights, and willingness to contribute have been instrumental to the success of this research.

## SUPPLEMENTARY FIGURE LEGENDS

**Figure S1(A).**
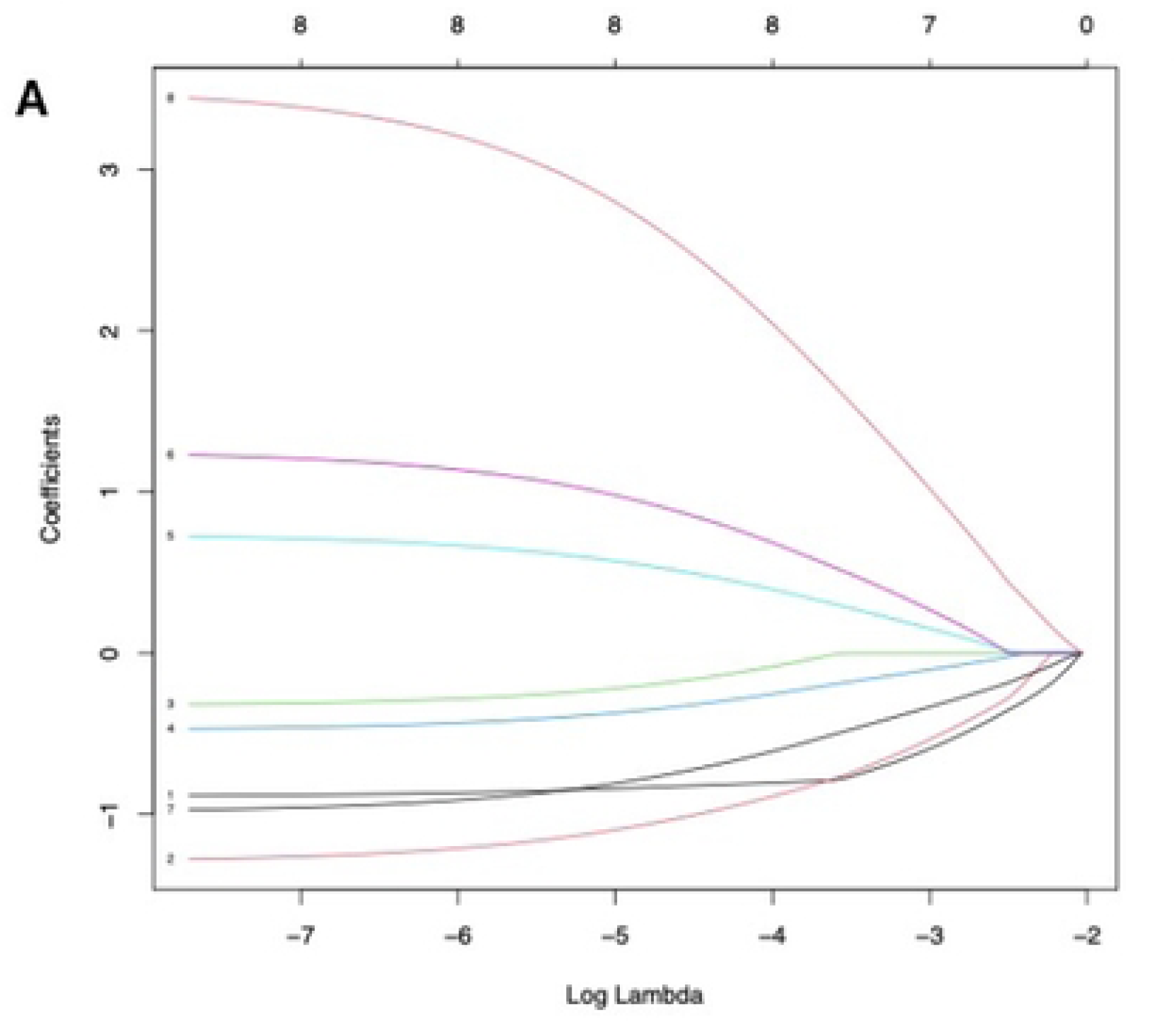
LASSO coefficient trajectories across varying lambda (λ) values. This plot illustrates how the estimated coefficients of predictors change as the regularization parameter λ increases. As λ grows, the model imposes stronger penalization, driving more coefficients to zero and resulting in increasingly sparse models that retain only the most influential predictors.

**Figure S1(B).**
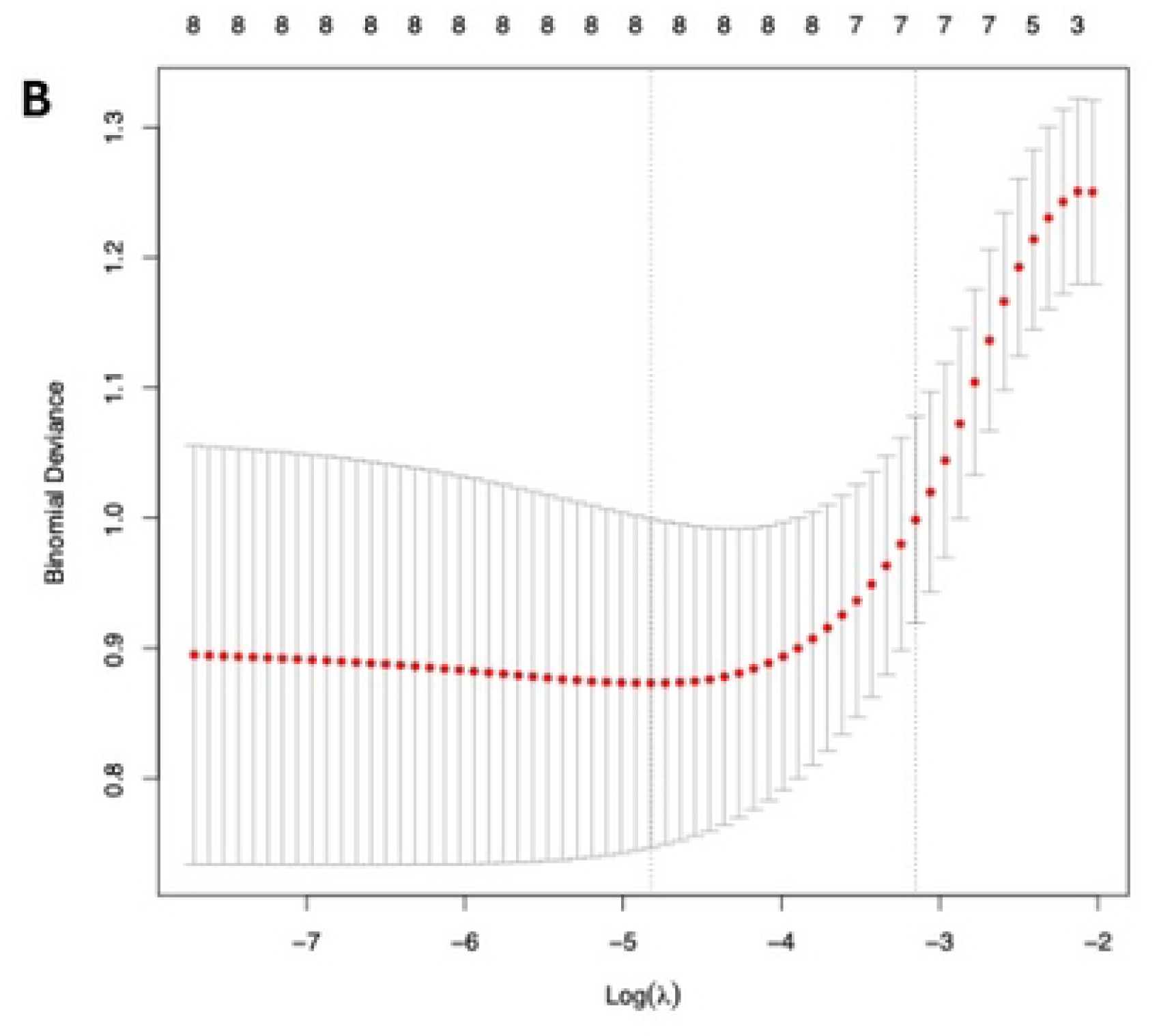
Cross-validation curve for LASSO regression on the training dataset. The plot shows the 10-fold cross-validated binomial deviance as a function of log-transformed lambda (λ), the regularization tuning parameter. This visualization supports model optimization by identifying the λ value that minimizes deviance, thereby selecting the most effective level of sparsity for the LASSO-regularized model.

**Figure S2(A).**
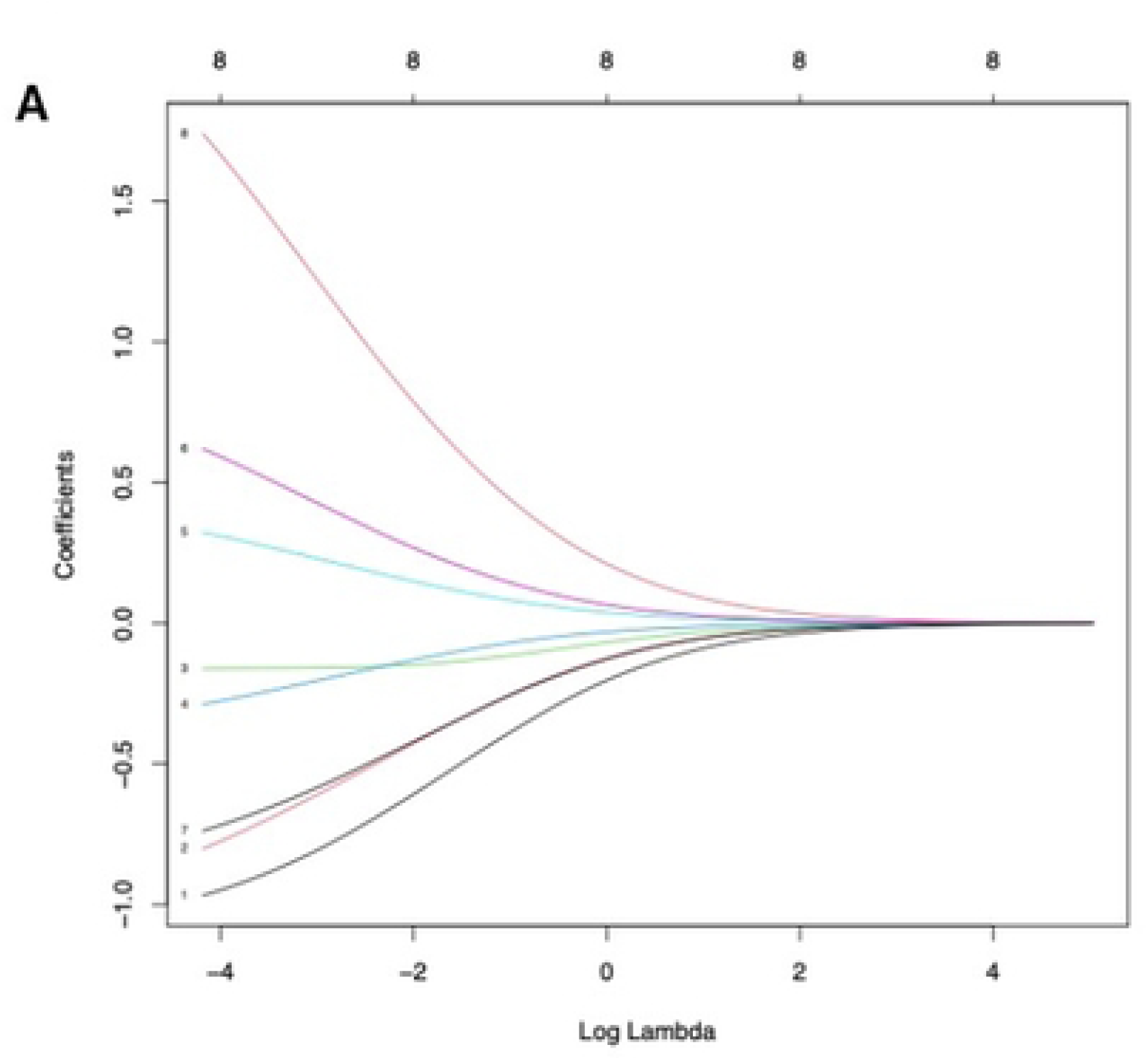
Ridge regression coefficient paths across varying lambda (λ) values. This plot illustrates how the estimated coefficients of predictors evolve as the regularization parameter λ increases. As λ grows, the model increasingly penalizes complexity, causing the coefficients to shrink toward zero—resulting in progressively sparser models with fewer influential predictors.

**Figure S2(B).**
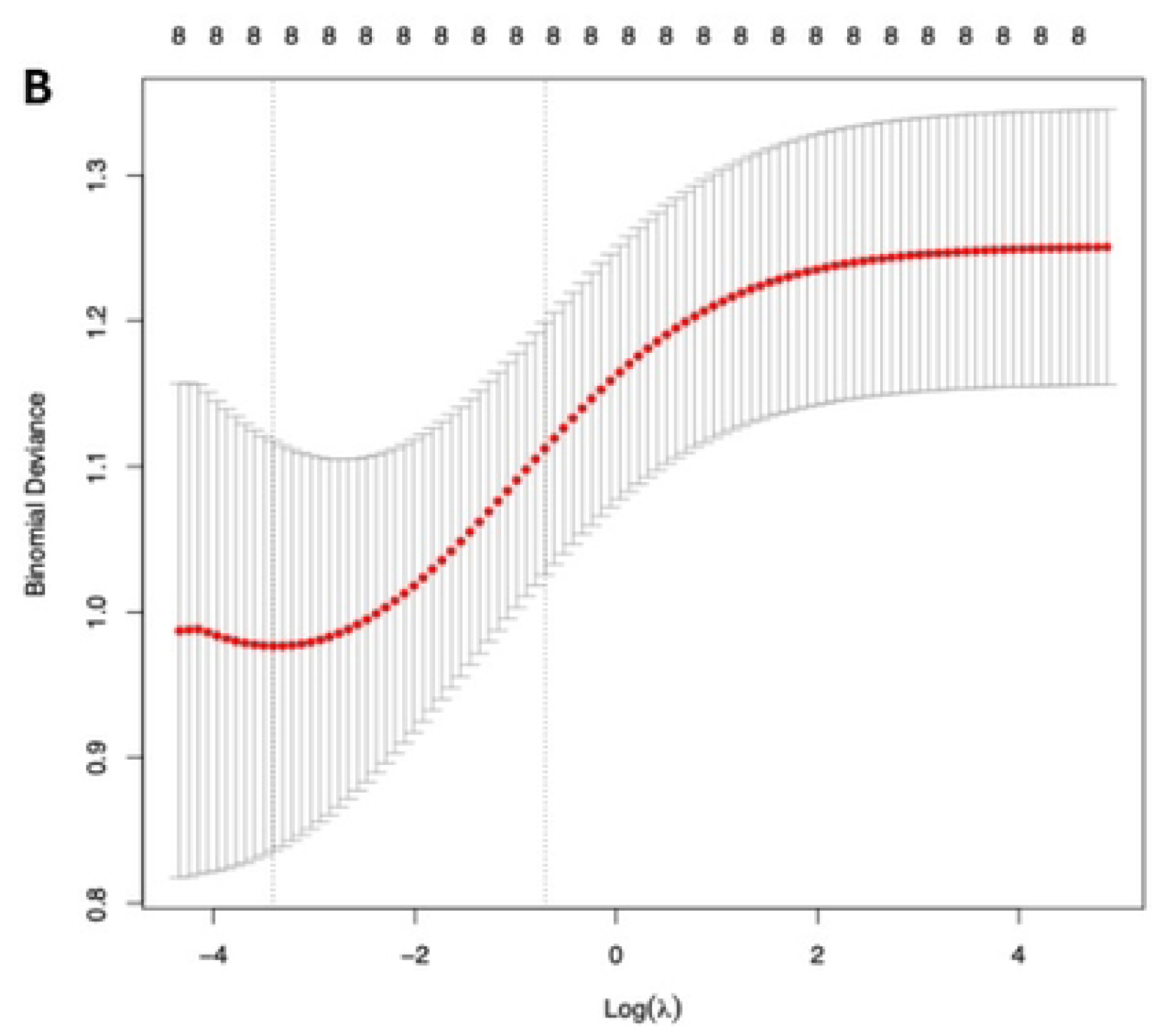
Cross-validation for ridge regression on the training dataset. The plot displays the 10-fold cross-validated binomial deviance as a function of log-transformed lambda (λ), the regularization tuning parameter. Although labeled for ridge regression, the curve reflects performance metrics for a lasso-regularized model, aiding in the selection of the optimal λ that minimizes deviance and enhances model generalization.

**Figure S3.**
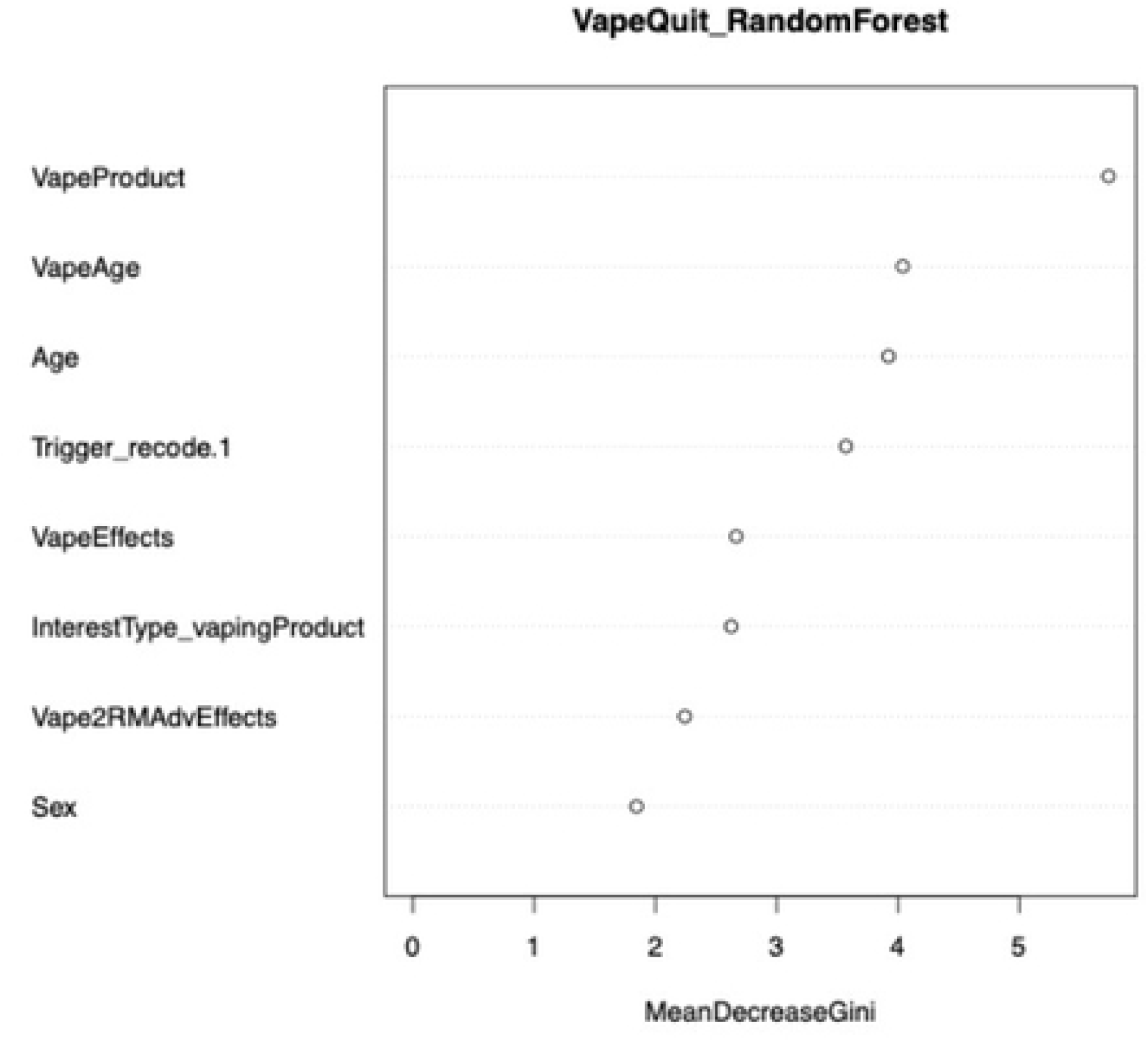
Variable Importance Plot from Random Forest Model. The plot displays the relative importance of predictor variables based on Mean Decrease Accuracy and Mean Decrease Gini. Variables are ranked from most to least important, with higher values indicating greater contribution to model performance

